# Pregnancy preparation amongst women and their partners in the UK; how common is it and what do people do?

**DOI:** 10.1101/2022.12.19.22283057

**Authors:** Catherine Louise Stewart, Jennifer Anne Hall

## Abstract

**Background:** Pregnancy preparation, to establish a healthy lifestyle within the preconception period, has been shown to reduce adverse maternal and neonatal outcomes. Despite its importance, we know very little about if and how people prepare for pregnancy in the UK.

**Methods:** As part of the P3 study, women in the UK were invited to complete an online survey about pregnancy preferences, including the Desire to Avoid Pregnancy (DAP) Scale. 274 participants were currently trying, thinking, or maybe thinking about getting pregnant and were asked about pregnancy preparations. The changes that women, and their partners, made in preparation for pregnancy, reasons for not preparing, and associations with sociodemographics were investigated in univariate and multivariate analyses.

**Results:** Of the 274 women, less than half (n=134, 49%) reported making any changes in preparation for pregnancy, with the most common changes being “eating healthier” (55%) and “folicacid” (54%). The main reason for not preparing was “only thinking about getting pregnant” (38%). 92 women answered questions about partner preparations; only 24% of partners were preparing, with the most common changes being “eating healthier” (64%) and “reducing alcohol” (50%). The main reason for partners not preparing was “already healthy” (51%). DAP score was the only significant factor affecting pregnancy preparation; every one-point increase in DAP score reduced the odds of a woman preparing for pregnancy by 78% (OR 0.22, 95%CI 0.15-0.34).

**Conclusion:** Interventions addressing pregnancy preparation for women, and their partners, are needed. These strategies should target women thinking about pregnancy, to ensure the full benefits of preconception care are received.

## Introduction

Pregnancy preparation - establishing a healthy lifestyle within the preconception period - has been shown to reduce adverse maternal and neonatal outcomes, such as pregnancy loss, intrauterine growth restriction and low birthweight (1, 2). The health of both mother and father before pregnancy has also been shown to have long term impacts on the health of future generations, including the risk of obesity, coronary heart disease, type 2 diabetes and neurodevelopmental disorders (3, 4).

Currently over 90% of women of reproductive age have at least one behavioural or medical risk factor before pregnancy, (5). Almost 50% of women in the UK are either overweight (BMI >25 kg/m^2)^ or obese (BMI >30 kg/m^2^) when they become pregnant (1, 6) which is associated with substantial risks for both mother and child, including sub-fertility, pregnancy complications, congenital fetal malformations and stillbirth (1, 7-9). However, it is not just maternal obesity that is an issue; paternal obesity has also been linked to impaired fertility, increased risk of adverse pregnancy outcomes and chronic disease risk in offspring (1, 10, 11). Despite the prevalence of overweight and obesity, micronutrient deficiencies continue to be a growing issue globally; studies have shown that many women of reproductive age will have dietary intakes below the recommend reference nutrient intake (RNI) for many micronutrients (1). Micronutrient deficiency during pregnancy can cause numerous adverse effects. For example, folic acid deficiency is associated with neural tube defects and other congenital abnormalities (12), iodine deficiency can adversely affect cognitive development (13, 14) and iron deficiency can increase the risk of impaired cognitive development in offspring, with studies suggesting that supplementation during pregnancy does not reverse the effect, highlighting the importance of preconception supplementation (15-17). Data from the UK National Diet and Nutrition Survey indicated that almost 90% of women of childbearing age have a folate concentration below the recommended level for preventing neural tube defects (18).

Preconception care involves the identification of risk factors alongside health promotion and health intervention, with the aim of improving the health of woman, and their partners, prior to getting pregnant, to help minimise risks and reduce adverse pregnancy outcomes (19, 20). For example, aiming to achieve a healthy weight, through a healthy diet, can help reduce the risks associated with overweight and obesity as well as help improve some micronutrient deficiencies. For some micronutrients, such as folate, it can be hard to achieve adequate levels through diet alone, therefore micronutrient supplementation, or food fortification are important. Folic acid supplementation, for at least three months prior to pregnancy (through to 12 weeks of pregnancy), has been shown to be effective at decreasing the risk of pre-eclampsia, miscarriage and low-birth weight, in addition to reducing the risk of neural tube defects, by as much as 70% (1, 21, 22). Other lifestyle changes such as reducing or quitting smoking and alcohol intake can also significantly reduce the risk of adverse outcomes such as intrauterine growth restriction, premature birth, low birth weight and congenital malformations, as well as improving fertility (23-27).

The timing of the preconception period is a debated topic (28), but is often defined as three months prior to conception. However, time to conception is different for every couple and can only be calculated retrospectively. Furthermore, numerous health issues that affect pregnancy, such as obesity and smoking, often take time and support to be tackled effectively (5). As a result, it is recommended that people start preparing for pregnancy when they start *thinking* about having a baby, to ensure they have adequate time to receive the benefits of preconception care (1, 5, 29).

Despite the importance of preconception health, there appears to be, within the general public, a lack of awareness of preconception care and knowledge of the impact that poor preconception health can have (30, 31). There is also limited research into if and how people prepare for pregnancy. This paper aims to identify the changes that women, and their partners, make in preparation for pregnancy, the reasons for not preparing and the factors associated with pregnancy preparation.

## Methods

### Study Setting and Design

As part of the P3 study, women in the UK were invited to participate in an online survey about pregnancy preferences. The data collection methods for this study have been previously described in detail elsewhere (32). Briefly, data was collected from non-pregnant women, aged 15 or over, living in the UK who had not gone through the menopause or been sterilised. Women completed a baseline survey online and were then invited to complete the survey again every three months for one year, unless they had an ongoing pregnancy at two consecutive time points. Overall, 994 women completed the baseline survey and almost 90% of eligible participants completed follow-up at 12 months; those lost to follow-up were not significantly different on key socio-demographic factors (32).

Of the 994 women who took part in the baseline survey, 274 women were asked about pregnancy preparation; these were women who answered “Yes” to currently trying to get pregnant, “Yes” or “Maybe” to thinking about getting pregnant in the next year, or “No” to using contraception in the last 30 days because they were either “Trying to get pregnant” or “Didn’t mind if they got pregnant” (Figure 1). 92 of the 994 women were asked about partner preparations; these were women who answered “No” to using contraception in the last 30 days because they were either “Trying to get pregnant” or “Didn’t mind if they got pregnant”.

**Figure 1.**
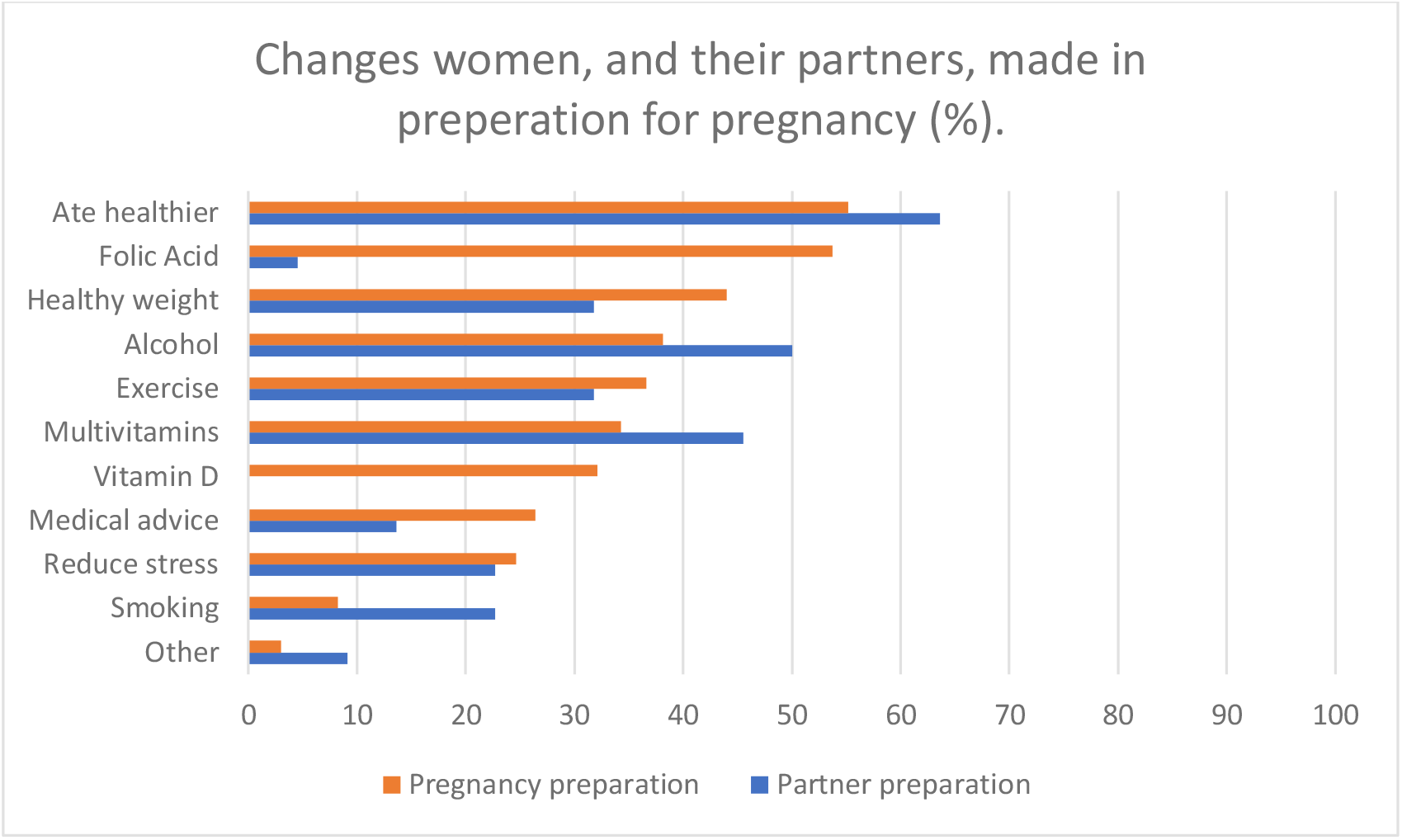
The preparations that women, and their partners, made in preparation for pregnancy (%).

Women were asked if they (and their partner) had done anything in preparation for pregnancy, if yes there were asked what changes they had made and if no, they were asked why they had not made any changes. All questions had multiple-choice answers with ‘other’ as an option. The changes that women, and their partners, made in preparation for pregnancy, as well as the reasons given for not preparing, were investigated. Both univariate and multivariate analyses were then performed to determine factors associated with pregnancy preparation and partner preparation.

### Measures

#### Pregnancy preparation

We created a binary variable of pregnancy preparation (yes/no) and a binary variable of partner preparation (yes/no). These outcome variables were derived from the answers to two, self-reported questions in the baseline survey; “Are you doing anything to improve your health in preparation for pregnancy?” and “Is your partner doing anything in preparation for pregnancy?”.

Women were categorised into one of four pregnancy intention groups (‘trying’, ‘thinking’, ‘maybe thinking’ or ‘not trying or thinking”) based on their answers to the ‘trying’ (‘are you currently trying to get pregnant?”) or ‘thinking’ (‘are you thinking about getting pregnant in the next year?’) questions. Women in the ‘trying’, ‘thinking’, or ‘maybe thinking’ groups were asked about pregnancy preparations.

While the pregnancy preparation questions were asked to women based on their answers to the ‘trying’ and ‘thinking’ questions, within the statistical analysis we chose to use Desire to Avoid Pregnancy (DAP) score to measure pregnancy intention as it has been shown to be a better predictor of pregnancy (33).

#### Desire to Avoid Pregnancy (DAP) Scale

The Desire to Avoid Pregnancy (DAP) scale is a psychometrically validated measure of a person’s preferences about a potential future pregnancy. The DAP scale has been validated for use within the UK and shown to be highly predictive of pregnancy (32, 34). The DAP scale is a continuous measure, with scores ranging from 0 (no desire to avoid pregnancy) to 4 (high desire to avoid pregnancy).(32).

#### Statistical Analysis

To assess the determinants of pregnancy preparation, univariate ordered logistic regressions of the chosen variables were performed to investigate the relationship between the variable and pregnancy preparation. Multivariable ordered logistic regression was then used with the pregnancy preparation variable as the outcome measure. All variables, regardless of their significance in the univariate analysis, were included in the multivariable analysis. This was decided as most of the variables are correlated and therefore, while the variables may be insignificant on their own, there is potential for negative confounding, with relationships being revealed in the multivariable analysis. All variables were introduced simultaneously, and any variables found to be non-significant (p>0.05) were removed using manual backwards stepwise elimination, starting with the variable with the highest p-value. The relationship between the variables and partner preparations was investigated using the same strategy.

## Results

### Participant Characteristics

The socio-demographic characteristics of the 994 women who took part in the baseline survey have previously been published (32). Briefly, women ranged in age from 15-50 years, with a median age of 31, 83.9% were white and 68.9% were educated to at least undergraduate level. The majority described themselves as heterosexual (81.6%), 48.2% were married, and 56.3% had been pregnant at least once before. The socio-demographic characteristics of all women, as well as a breakdown by pregnancy intention, are shown in Table 1.

**Table 1.** Characteristics of all women, and the women within each pregnancy intention group (‘trying’, ‘thinking’, ‘maybe thinking’ and ‘not trying/thinking’).

| Variable | All woman<br>N = 994 | Trying<br>N=80<br>(8.2%) | Thinking<br>N = 96<br>(9.8%) | Maybe<br>Thinking<br>N = 97<br>(9.9%) | Not trying/<br>thinking<br>N = 707<br>(72.1%) | p-value |
| --- | --- | --- | --- | --- | --- | --- |
| **Age** |  |  |  |  |  |  |
| Mean (SD) | 29.7 (8.3) | 32.8 (5.4) | 32.4 (3.9) | 30.4 (6.1) | 28.9 (8.9) | **<0.001** |
| Range | 15-50 | 21-47 | 21-41 | 16-42 | 15-50 |  |
|  | **N (%)** |  |  |  |  |  |
| **Education** |  |  |  |  |  |  |
| Below undergraduate | 300 (31.2) | 13 (16.2) | 7 (7.6) | 17 (17.7) | 261 (37.9) | **<0.001** |
| Undergraduate or above | 663 (68.8) | 67 (83.8) | 85 (92.4) | 79 (82.3) | 427 (62.1) |  |
| **Relationship status** |  |  |  |  |  |  |
| No relationship | 152 (15.7) | 1 (1.3) | 1 (1.1) | 9 (9.4) | 138 (20.0) | **<0.001** |
| Relationship | 337 (34.7) | 19 (23.7) | 15 (15.8) | 33 (34.4) | 270 (39.0) |  |
| Married | 481 (49.6) | 60 (75.0) | 79 (83.1) | 54 (56.2) | 284 (41.0) |  |
| **Previous pregnancy** |  |  |  |  |  |  |
| 0 | 434 (43.8) | 22 (27.5) | 22 (22.9) | 34 (35.0) | 348 (49.4) | **<0.001** |
| 1 | 169 (17.0) | 23 (28.8) | 33 (34.4) | 32 (33.0) | 79 (11.2) |  |
| 2+ | 389 (39.2) | 35 (43.7) | 41 (42.7) | 31 (32.0) | 278 (39.4) |  |
| **Ethnicity** |  |  |  |  |  |  |
| White | 834 (86.0) | 68 (85.0) | 87 (91.6) | 80 (84.2) | 598 (85.6) | 0.531 |
| BAME | 136 (14.0) | 12(15.0) | 8 (8.4) | 15 (15.8) | 100 (14.4) |  |
| **Religious** |  |  |  |  |  |  |
| Yes | 373 (38.4) | 36 (45.0) | 41 (43.2) | 39 (40.6) | 643 (63.4) | 0.065 |
| No | 598 (61.6) | 44 (55.0) | 54 (56.8) | 57 (59.4) | 254 (36.6) |  |
| **English 1<sup>st</sup> Language** |  |  |  |  |  |  |
| Yes | 898 (92.5) | 76 (95.0) | 86 (90.5) | 86 (90.5) | 643 (92.8) | 0.675 |
| No | 73 (7.5) | 4 (5.0) | 9 (9.5) | 10 (9.5) | 50 (7.2) |  |

The average DAP score for each pregnancy intention group is shown in Table 2.

**Table 2.** The average DAP score for each of the four Pregnancy intention groups.

|  | Pregnancy intention |  |  |  | P Value |
| --- | --- | --- | --- | --- | --- |
|  | Trying | Thinking | Maybe thinking | Not trying/<br>thinking |  |
| **Average DAP Score (range)** | 0.82<br>(0 - 1.86) | 1.4<br>(0 - 2.79) | 1.8<br>(0.29 - 3.64) | 3.0<br>(0.93 – 4) | <0.001 |

### Pregnancy Preparations

Of the 274 women asked about pregnancy preparations, less than half (n=134, 49%) reported making any changes in preparation for pregnancy. Women who were preparing for pregnancy made between one and nine changes, with four being the average number of changes made by women in preparation for pregnancy. The most common changes made by women were “eating healthier” (n=74; 55%) and “taking folic acid” (n=72; 54%). All the changes made are shown in Figure 1.

### Partner preparations

92 women were asked questions about partner preparation; only 22 women reported their partner making any changes in preparation for pregnancy (24%), 67 women reported no changes (73%) and three women did not have a partner (3%) (these women were subsequently dropped from analysis). Partners made between one and seven changes in preparation for pregnancy, with three being the average number of changes made by partners. The most common change reported was “eating healthily” (n=14; 64%) followed by “stopping or cutting down alcohol” (n=11; 50%). The other changes made by partners in preparation for pregnancy are also shown in Figure 1.

### Reasons for not preparing

140 women reported making no changes in preparation for pregnancy, the reasons given for not preparing are shown in Figure 2. The most common reason given for not preparing was “right now, I’m only thinking about trying to get pregnant” (n=52, 38%), with other reasons including “there is nothing I need to do to improve my health in preparing for pregnancy because I am already healthy” (n=25, 18%) and “once I am pregnant, I will take some action for a healthy pregnancy” (n=21, 15%). The reasons given for why partners were not preparing are also show in in Figure 2; the main reasons were “there is nothing my partner needs to do to improve their health in preparation for pregnancy because they are already healthy” (n=34; 51%) and “we did not know there was anything my partner should be doing before pregnancy” (n=13; 19%).

**Figure 2.**
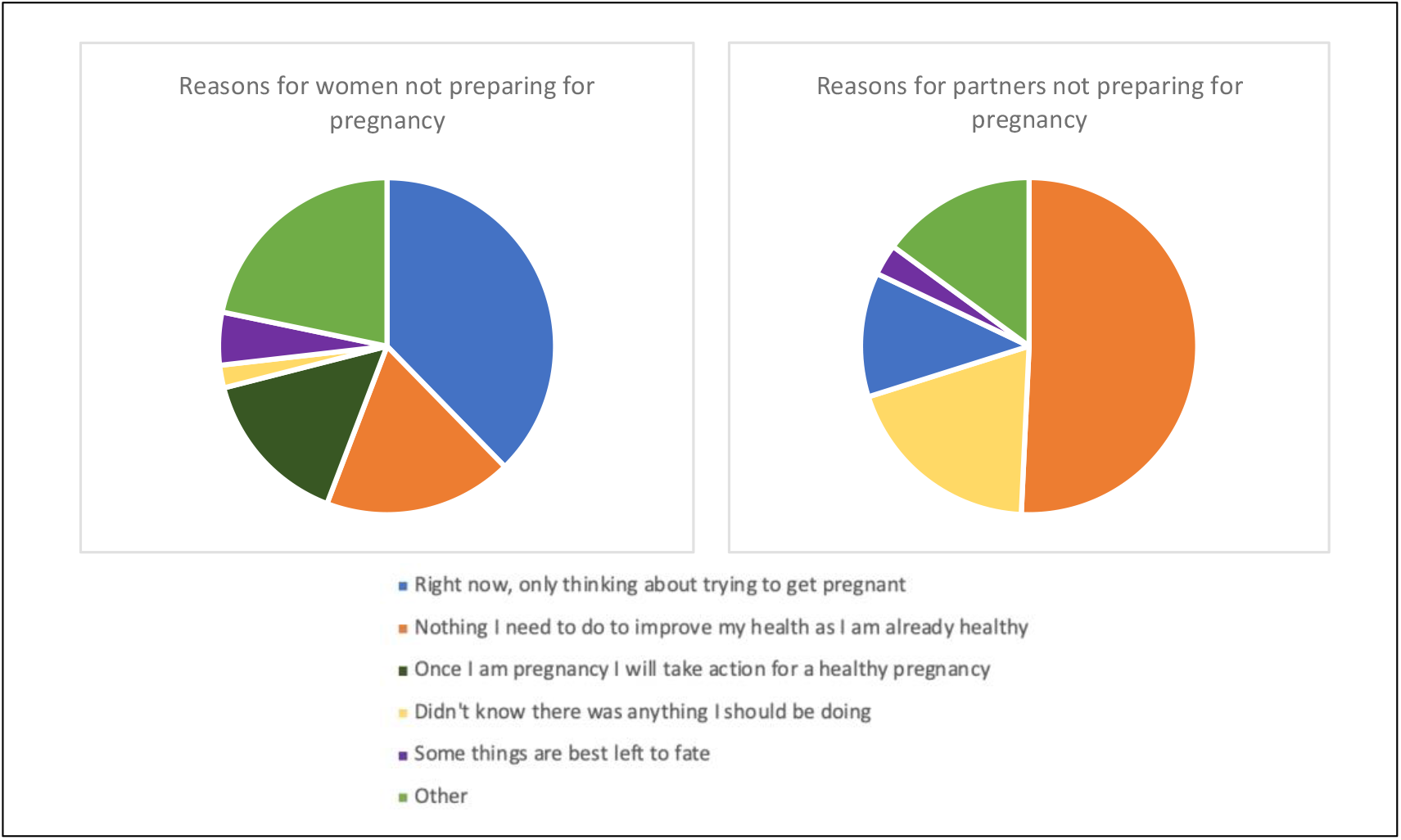
Reasons given for why women and their partners were not preparing for pregnancy.

### Pregnancy preparation analysis

Univariate analysis showed that relationship status and DAP score both had a significant effect on pregnancy preparation; married women were over five-times more likely to prepare for pregnancy than women not in a relationship (OR 5.6, 95%CI 1.2-26.25). All other variables - age, education, previous pregnancy, ethnicity, religion and first language - were not significant (Table 3).

**Table 3.** Univariate and multivariable linear regressions of characteristics associated with pregnancy preparation.

| Variable | Univariate analysis |  |  | Multivariate analysis |  |  |
| --- | --- | --- | --- | --- | --- | --- |
|  | OR | 95% CI | P-value | OR | 95% CI | P-value |
| **Age, years** | 1.03 | 0.98 – 1.08 | 0.262 |  |  |  |
| **Education** | Below degree level as baseline |  |  |  |  |  |
| Undergraduate degree or above | 1.05 | 0.53 – 2.08 | 0.898 |  |  |  |
| **Relationship status** | No relationship as baseline |  |  |  |  |  |
| Relationship | 3.82 | 0.78 – 18.72 | **0.047** |  |  |  |
| Married | 5.60 | 1.2 – 26.25 |  |  |  |  |
| Previous pregnancy | 0 as baseline |  |  |  |  |  |
| 1 | 0.85 | 0.46 – 1.56 | 0.861 |  |  |  |
| 2+ | 0.96 | 0.53 – 1.71 |  |  |  |  |
| Ethnicity | White as baseline |  |  |  |  |  |
| BAME | 0.98 | 0.48 – 1.99 | 0.949 |  |  |  |
| Religious | No as baseline |  |  |  |  |  |
| Yes | 1.29 | 0.80 – 2.09 | 0.294 |  |  |  |
| First language | No as baseline |  |  |  |  |  |
| Yes | 1.27 | 0.54 – 3.00 | 0.588 |  |  |  |
| DAP | 0.22 | 0.15 – 0.34 | <0.001 | 0.22 | 0.15 – 0.34 | <0.001 |

Following multivariable analysis, DAP score was the only variable found to have a significant effect on pregnancy preparation (Table 3), with each increasing DAP point (range: 0–4) resulting in a 78% decrease in the odds of a woman preparing for pregnancy (OR 0.22, 95%CI 0.15-0.34), i.e., as a women’s desire to avoid pregnancy increases, the chances of her preparing for pregnancy significantly decreases.

### Partner preparation analysis

DAP score was shown to be the only factor significantly associated with partner preparation following both univariate and multivariate analysis (Table 4); for every one-point increase in DAP score, the odds of a woman’s partner preparing for pregnancy decreased by 87% (OR 0.13, 95%CI 0.03-0.46).

**Table 4.** Univariate and multivariable linear regressions of characteristics associated with partner preparation.

|  | Univariate analysis |  |  | Multivariate analysis |  |  |
| --- | --- | --- | --- | --- | --- | --- |
| Variable | OR | 95% CI | P-value | OR | 95% CI | P-value |
| Age, years | 0.91 | 0.82 – 1.01 | 0.063 |  |  |  |
| Education | Below degree level as baseline |  |  |  |  |  |
| Undergraduate degree or above | 0.51 | 0.15 – 1.72 | 0.275 |  |  |  |
| Relationship status | Not married as baseline |  |  |  |  |  |
| Married | 0.52 | 0.17 – 1.52 | 0.231 |  |  |  |
| Previous pregnancy | 0 as baseline |  |  |  |  |  |
| 1 | 0.47 | 0.13 – 1.72 | 0.431 |  |  |  |
| 2+ | 0.53 | 0.17 – 1.64 |  |  |  |  |
| Ethnicity | White as baseline |  |  |  |  |  |
| BAME | 0.27 | 0.03 – 2.25 | 0.227 |  |  |  |
| Religion belong | No as baseline |  |  |  |  |  |
| Yes | 0.63 | 0.23 – 1.69 | 0.353 |  |  |  |
| First language | No as baseline |  |  |  |  |  |
| Yes | 0.98 | 0.10 – 9.98 | 0.989 |  |  |  |
| DAP | 0.13 | 0.03 – 0.48 | 0.002 | 0.13 | 0.03 – 0.48 | 0.002 |

## Discussion

Overall, there appears, within the UK, to be a lack of awareness amongst the general population of the importance of preconception health. This is not only shown by the inadequate number of people preparing for pregnancy - less than half of women (and less than a quarter of partners) reported making any changes - but is also exposed in the reasons given for not preparing. When women were asked why they were not preparing for pregnancy the most common reasons given were “right now, I’m only think about trying to get pregnant” and “once I am pregnant, I will take some action for a healthy pregnancy”.

This highlights a lack of knowledge of the time that preparing for pregnancy can take and the impact that it can have; not only on pregnancy outcomes and the future health of offspring, but also the chances of getting pregnant (1, 3, 7, 25, 26). Furthermore, research has shown that intervening during pregnancy is often too late and therefore preconception preparations are vital (15, 35). Another common reason given for not preparing was “there is nothing I need to do to prepare for pregnancy as I am already healthy”. While this may be true (although previous research has shown people often overestimate their own health status (2)) every woman, regardless of their health, should be taking (at least) 400mg of folic acid for at least three months prior to pregnancy to reduce the risk of neural tube defects (36, 37). Despite this recommendation, almost 50% of women who were preparing for pregnancy were not taking folic acid, indicating a lack of understanding of the importance of preconception folic acid. Additionally, one of the main reasons given for why partners were not preparing was “We didn’t know that they should be doing anything” suggesting that people are unaware of the impact that preconception paternal health can have (4, 10, 11).

DAP score was shown to be the only factor significantly associated with pregnancy preparation; as a woman’s desire to avoid pregnancy increased the odds of her preparing for pregnancy decreased. This is in line with previous research showing that pregnancy intention is linked to maternal health behaviours (38, 39). The DAP scale has been shown to provide nuance that is often missed by a single question (33). For example, women who categorised themselves as only thinking about pregnancy within the next year, had an average DAP score of 1.4 (indicating a relatively low desire to avoid pregnancy); previous work on the predictive ability of the DAP suggests that around 43% of these women would become pregnant within 12 months (34). Furthermore, almost 30% of the women in the ‘thinking’ group were not using any contraception. Therefore, while they may only be thinking about getting pregnant, they still have a relatively high chance of pregnancy in the near future. In addition, it is recommended that women, and their partners, start to prepare for pregnancy before actively trying to conceive (so within the thinking period) to ensure they receive the full benefits of preconception care (1, 5). Consequently, women who are thinking about pregnancy make up a critical group and it will be vital to increase the number of women within this group preparing for pregnancy, in order to improve pregnancy outcomes.

The lack of awareness and low uptake of pregnancy preparations shown in this study fits with published literature (2, 30, 31), highlighting the need for strategies aimed at improving preconception knowledge. The results of this study support previous research, stressing the need for normalising conversations around pregnancy intention and pregnancy preparations, suggesting that clinicians should routinely ask people of reproductive age questions about pregnancy intentions(5, 33). Although the DAP scale has been shown to provide more nuance than other measures, it is made up of 14 questions which can limit its clinical use. Therefore, until there is further research into the clinical applications of the DAP, for example as a digital tool or using a single DAP question or combination of DAP questions (33), clinicians should routinely ask people of reproductive age questions such as “Are you thinking about pregnancy in the next year?”(5, 40) or ‘Can I help you with any reproductive health services today?’(41, 42). Based on the response clinicians should discuss preconception health with individuals who are thinking about pregnancy or contraception with those who wish to avoid pregnancy. Finally, this study also highlights the importance of including partners in these conversations, to ensure people know how impactful paternal health can be.

### Strengths and Limitations

The analysis used data from a large, broadly representative dataset, suggesting generalisability of our findings. However, within the P3 Study questionnaire, the questions on pregnancy preparation were about *changes* made in preparation for pregnancy. As a result, the data collected gives us a guide to the relative likelihood of the changes women, and their partners, make in preparation for pregnancy, but does not give us the absolute prevalence of those changes within the UK. Furthermore, we were unable to analyse partner preparation by sex of partner, as there were too small numbers.

## Conclusion

Overall, the results of this study show a lack of pregnancy preparation, likely due to a lack of knowledge, among women and their partners, of the importance of preconception care and the impact that poor preconception health can have on pregnancy outcomes and the future health of offspring. This highlights the need for strategies aimed at increasing the awareness of the importance of health before pregnancy within the UK. These strategies should be focused on people who are thinking about getting pregnant (not just those who are actively trying), to ensure that they have enough time to properly prepare for pregnancy, enabling them to receive the full benefits of better preconception health.

## Supporting information

STROBE Checklist

## Data Availability

All data produced are available online at the UCL Research Data Repository.

## Acknowledgements

We would like to thank all the women who took part in the P3 study.

## Data availability statement

The dataset is available in the UCL Research Data Repository.

## References

1. Stephenson J, Heslehurst N, Hall J, Schoenaker DAJM, Hutchinson J, Cade JE, et al. Before the beginning: nutrition and lifestyle in the preconception period and its importance for future health. The Lancet. 2018;391(10132):1830–41.

2. Maas VYF, Poels M, Kievit MHd, Hartog AP, Franx A, Koster MPH. Planning is not equivalent to preparing, how Dutch women perceive their pregnancy planning in relation to preconceptional lifestyle behaviour change-a cross-sectional study. BMC pregnancy and childbirth. 2022;22(1):577.

3. Godfrey KM, Reynolds RM, Prescott SL, Nyirenda M, Jaddoe VW, Eriksson JG, et al. Influence of maternal obesity on the long-term health of offspring. Lancet Diabetes Endocrinol. 2017;5(1):53–64.

4. Fleming TP, Watkins AJ, Velazquez MA, Mathers JC, Prentice AM, Stephenson J, et al. Origins of lifetime health around the time of conception: causes and consequences. Lancet. 2018;391(10132):1842–52.

5. Stephenson J, Schoenaker DA, Hinton W, Poston L, Barker M, Alwan NA, et al. A wake-up call for preconception health: a clinical review. Br J Gen Pract. 2021;71(706):233–6.

6. NHS. Statistics on Obesity, Physical Activity and Diet 2020 [2 November 2022]. Available from: https://digital.nhs.uk/data-and-information/publications/statistical/statistics-on-obesity-physical-activity-and-diet/england-2020/part-3-adult-obesity-copy.

7. Poston L, Caleyachetty R, Cnattingius S, Corvalán C, Uauy R, Herring S, et al. Preconceptional and maternal obesity: epidemiology and health consequences. Lancet Diabetes & Endocrinology. 2016;4(12):1025–36.

8. Law DCG, Maclehose RF, Longnecker MP. Obesity and Time to Pregnancy. Hum Reproduction. 2007;22(2):414–20.

9. Marchi J, Berg M, Dencker A, Olander EK, Begley C. Risks associated with obesity in pregnancy, for the mother and baby: a systematic review of reviews. Obesity Reviews. 2015;16(8):621–38.

10. Hieronimus B, Ensenauer R. Influence of maternal and paternal pre-conception overweight/obesity on offspring outcomes and strategies for prevention. European Journal of Clinical Nutrition volume. 2021;75:1735–44.

11. Lin J, Gu W, Huang H. Effects of Paternal Obesity on Fetal Development and Pregnancy Complications: A Prospective Clinical Cohort Study. Frontiers in Endocrinology. 2022;13.

12. Greenberg J, Bell S, Guan Y, Yu Y. Folic Acid supplementation and pregnancy: more than just neural tube defect prevention. Reviews in Obstetrics and Gynecology. 2011;4(2):52–9.

13. Zimmermann MB. Iodine deficiency in pregnancy and the effects of maternal iodine supplementation on the offspring: a review. Am J Clin Nutr. 2009;89(2):668s–72s.

14. Pearce EN. Monitoring and effects of iodine deficiency in pregnancy: still an unsolved problem? European Journal of Clinical Nutrition. 2013;67(5):481–4.

15. Janbek J, Sarki M, Specht IO, Heitmann BL. A systematic literature review of the relation between iron status/anemia in pregnancy and offspring neurodevelopment. European Journal of Clinical Nutrition. 2019;73(12):1561–78.

16. Means RT. Iron Deficiency and Iron Deficiency Anemia: Implications and Impact in Pregnancy, Fetal Development, and Early Childhood Parameters. Nutrients. 2020;12(2).

17. Bath SC, Steer CD, Golding J, Emmett P, Rayman MP. Effect of inadequate iodine status in UK pregnant women on cognitive outcomes in their children: results from the Avon Longitudinal Study of Parents and Children (ALSPAC). Lancet. 2013;382(9889):331–7.

18. PHE. National Diet and Nutrition Survey: results from years 9 to 11 (2016 to 2017 and 2018 to 2019). 2020 [3 November 2022]. Available from: https://www.gov.uk/government/statistics/ndns-results-from-years-9-to-11-2016-to-2017-and-2018-to-2019.

19. Ignaszak-Kaus N, Ozegowska K, Piekarski P, Pawelczyk L, Jędrzejczak P. Planning and preparation for pregnancy among women with and without a history of infertility. Ginekol Pol. 2018;89(2):74–9.

20. Bortolus R, Oprandi NC, Rech Morassutti F, Marchetto L, Filippini F, Agricola E, et al. Why women do not ask for information on preconception health? A qualitative study. BMC Pregnancy Childbirth. 2017;17(1):5.

21. De-Regil LM, Peña-Rosas JP, Fernández-Gaxiola AC, Rayco-Solon. P. Effects and safety of periconceptional oral folate supplementation for preventing birth defects. Cochrane database of systematic reviews. 2015;12.

22. He Y, Pan A, Hu FB, Ma X. Folic acid supplementation, birth defects, and adverse pregnancy outcomes in Chinese women: a population-based mega-cohort study. The Lancet. 2016;388:S91.

23. Lassi ZS, Imam AM, Dean SV, Bhutta ZA. Preconception care: caffeine, smoking, alcohol, drugs and other environmental chemical/radiation exposure. Reproductive Health. 2014;11(3):S6.

24. Li J, Qiu J, Lv L, Mao B, Huang L, Yang T, et al. Paternal factors and adverse birth outcomes in Lanzhou, China. BMC Pregnancy Childbirth. 2021;21(1):19.

25. de Angelis C, Nardone A, Garifalos F, Pivonello C, Sansone A, Conforti A, et al. Smoke, alcohol and drug addiction and female fertility. Reproductive Biology and Endocrinology. 2020;18(1):21.

26. Sharma R, Biedenharn KR, Fedor JM, Agarwal A. Lifestyle factors and reproductive health: taking control of your fertility. Reproductive Biology and Endocrinology. 2013;11(1):66.

27. Oostingh EC, Hall J, Koster MPH, Grace B, Jauniaux E, Steegers-Theunissen RPM. The impact of maternal lifestyle factors on periconception outcomes: a systematic review of observational studies. Reprod Biomed Online. 2019;38(1):77-94.n

28. Hill B, Hall J, Skouteris H, Currie S. Defining preconception: exploring the concept of a preconception population. BMC Pregnancy and Childbirth. 2020;20(1):280.s

29. CDC. Planning for Pregnancy 2022 [Available from: https://www.cdc.gov/preconception/planning.html.

30. Daly MP, White J, Sanders J, Kipping RR. Women’s knowledge, attitudes and views of preconception health and intervention delivery methods: a cross-sectional survey. BMC Pregnancy Childbirth. 2022;22(1):729.

31. McGowan L, Lennon-Caughey E, Chun C, McKinley MC, Woodside JV. Exploring preconception health beliefs amongst adults of childbearing age in the UK: a qualitative analysis. BMC Pregnancy Childbirth. 2020;20(1):41.

32. Hall J, Barrett G, Rocca C. Evaluation of the Desire to Avoid Pregnancy Scale in the UK: a psychometric analysis including predicitive validity. BMJ Open. 2022;12(7).

33. Hall J, Barrett G, Stephenson J, Edelman N, Rocca CH. The Desire to Avoid Pregnancy Scale: clinical considerations and comparison with other questions about pregnancy preferences. medRxiv. 2022:2022.11.07.22281888.

34. Hall J, Barrett G, Stephenson J, Rocca C, Edelman N. Predictive Ability of the Desire to Avoid Pregnancy Scale. medRxiv. 2022:2022.10.17.22281028.

35. Dodd JM, Louise J, Deussen AR, Grivell RM, Dekker G, McPhee AJ, et al. Effect of metformin in addition to dietary and lifestyle advice for pregnant women who are overweight or obese: the GRoW randomised, double-blind, placebo-controlled trial. The Lancet Diabetes & Endocrinology. 2019;7(1):15–24.

36. Mastroiacovo P, Leoncini E. More folic acid, the five questions: why, who, when, how much, and how. Biofactors. 2011;37(4):272–9.

37. De-Regil LM, Peña-Rosas JP, Fernández-Gaxiola AC, Rayco-Solon P. Effects and safety of periconceptional oral folate supplementation for preventing birth defects. Cochrane Database Syst Rev. 2015;2015(12):Cd007950.

38. Chuang CH, Hillemeier MM, Dyer AM, Weisman CS. The relationship between pregnancy intention and preconception health behaviors. Prev Med. 2011;53(1-2):85–8.

39. Chatterjee E, Sennott C. Fertility intentions and maternal health behaviour during and after pregnancy. Popul Stud (Camb). 2020;74(1):55–74.

40. Hammarberg K, Hassard J, de Silva R, Johnson L. Acceptability of screening for pregnancy intention in general practice: a population survey of people of reproductive age. BMC Family Practice. 2020;21(1):40.

41. Jones HE, Calixte C, Manze M, Perlman M, Rubin S, Roberts L, et al. Primary care patients’ preferences for reproductive health service needs assessment and service availability in New York Federally Qualified Health Centers. Contraception. 2020;101(4):226–30.

42. Manze MG, Romero DR, Sumberg A, Gagnon M, Roberts L, Jones H. Women’s Perspectives on Reproductive Health Services in Primary Care. 2020.

